# From surveillance maturity to analytical readiness: an estimand-first framework for real-time outbreak analysis under imperfect data

**DOI:** 10.64898/2026.08.12.26360299

**Authors:** Johan G. L. Verheyden, Celestin Nzanzu Mudogo, Wolfgang Jacquet

**Author notes:** Corresponding author: Celestin Nzanzu Mudogo. Submitting author (manuscript correspondence and portal submission): Johan G. L. Verheyden.

## Abstract

**Background:** Real-time outbreak analyses are often requested before surveillance systems have stabilised or epidemics have generated enough information for the desired inference. Existing approaches address surveillance quality, forecasting, estimands and identifiability separately, but do not provide a common rule for deciding which analytical product is supportable at a particular data vintage. We developed an estimand-first framework for analytical readiness.

**Methods:** The framework distinguishes surveillance maturity (S), epidemic-process informativeness (E) and estimand-specific analytical readiness (Rθ,v), defined as whether the available data vintage, observation process, method and decision-matched validation support a specified inference for a specified decision. We stress-tested four implications using longitudinal data from the 2018–2020 Ebola response in eastern Democratic Republic of the Congo (DRC), archived geographic forecasts, independent forecasting data from Western Area, Sierra Leone, and a targeted mortality-identifiability experiment.

**Results:** During a documented DRC surveillance disruption and recovery, seven-day persistence forecasts had all-health-zone absolute errors of 1, 3, 18 and 4 cases across pre-shock, acute-shock, early-recovery and recovery origins; the largest error occurred during early recovery. Four-week reported-case trend multipliers changed from 0.67 and 0.73 to 1.12 and 1.29, while the final fit was strongly overdispersed (Pearson dispersion 7.65), demonstrating asynchronous readiness across estimands. Archived geographic forecasts improved a Top-3 allocation decision over cumulative burden at only one origin despite consistently lower Brier scores for one specification. In Western Area, persistence forecast mean absolute error increased from 39.1 cases at one week to 82.3 at four weeks, and a history-to-horizon ratio did not define a universal threshold. An observed reported case-fatality ratio of 0.40 was compatible with constructed latent fatality values from 0.10 to 0.80; increasing the reported denominator narrowed sampling uncertainty without reducing structural uncertainty.

**Conclusions:** Analytical readiness is task- and vintage-specific rather than a property of a dataset. More data, model convergence or narrow intervals cannot substitute for estimand definition, observation-process awareness, decision-matched validation and explicit identification analysis.

## Background

Real-time outbreak analysis is conducted under an unavoidable asymmetry. Decisions about surveillance deployment, diagnostic capacity, clinical resources, geographic pre-positioning and risk communication must often be made early, when the information required to support those decisions is least complete. A case series may become available within days of outbreak recognition, while the system producing it is still expanding case finding, decentralising testing, clearing laboratory backlogs, revising classifications, reconciling geographic assignments and retrospectively incorporating earlier events. At the same time, the epidemic itself may not yet have generated enough observations to identify growth dynamics, epidemic curvature, geographic spread, clinical outcomes or competing transmission histories reliably.

These limitations are often treated collectively as a problem of “poor data”, but they arise from two fundamentally different sources. One concerns the observation system: whether surveillance processes are consistently detecting and reporting disease activity. The other concerns the epidemic process: whether enough epidemiological information has accumulated to support the parameter, prediction or comparison being requested. A technically effective surveillance system can observe an epidemic that remains too young for stable forecasting or model discrimination. Conversely, substantial transmission may already have occurred while an incomplete or rapidly changing surveillance system distorts when, where and in whom that transmission becomes visible.

This distinction matters because routinely reported outbreak data are not direct measurements of the latent epidemic. Reported cases are conditioned on case recognition, health-seeking behaviour, access, eligibility for testing, specimen collection, laboratory throughput, classification and reporting. Reported deaths introduce additional processes, including outcome follow-up, recognition of community deaths, retrospective confirmation and linkage of outcomes to previously reported cases. Reporting dates may therefore represent administrative recognition rather than dates of infection, symptom onset, diagnosis or death. Established work on emerging-epidemic inference has shown that incomplete observation and epidemic growth can interact to bias epidemiological estimates even where standard models appear statistically well behaved [1].

Several methodological traditions address parts of this problem. Nowcasting and reporting-delay models seek to reconstruct events that have occurred but have not yet entered surveillance records [2]. Public-health surveillance frameworks evaluate attributes including sensitivity, timeliness, data quality, representativeness and stability [3]. Epidemic-growth methods examine how much can be inferred from early trajectories and explicitly recognise departures from simple exponential dynamics [4]. Forecast-evaluation methods emphasise out-of-sample assessment rather than fit to a completed epidemic, while probabilistic forecasts require evaluation using proper scoring rules that reward both calibration and sharpness [5–7]. Retrospective evaluations of Ebola forecasting have similarly demonstrated the importance of reproducing the information that would have been available at each forecast date rather than judging a model principally by its fit to the completed epidemic [8].

These approaches solve important but different problems. Correcting a reporting delay does not establish that the upstream surveillance system generating that delay has become stable. Demonstrating computational convergence does not establish that the epidemiological parameter being estimated is identifiable. A model that predicts the centre of a national epidemic trajectory accurately may nevertheless provide poorly calibrated uncertainty. A spatial model may perform usefully in identifying a small set of locations requiring attention even when its absolute risk probabilities remain insufficiently calibrated. Likewise, stabilising an aggregate deaths-to-cases ratio does not transform it into a patient-level probability of death when outcomes, ascertainment and reporting processes remain unresolved. Analyses of Ebola outbreaks have repeatedly shown that case-fatality estimates are sensitive to the composition and completeness of available case and outcome data [9].

Existing frameworks thus address surveillance-system performance, target estimands, predictive validation and parameter identifiability largely as separate methodological problems. What is missing is an operational framework for determining, at a particular surveillance vintage, which kind of analytical product is supportable for a particular estimand and decision. Analytical readiness fills this gap by linking observation-system evidence, epidemic-process information, method-specific validation and identifiability to the strongest permissible inference.

The framework begins by separating surveillance maturity, an observation-system property, from epidemic-process informativeness, which concerns the accumulation of estimand-relevant epidemic signal. The present study asks a further question: what inference is supportable for a particular estimand at a particular data vintage?

That distinction nevertheless leaves a further methodological problem unresolved. Analytical readiness is not uniform across estimands. The information required to estimate early epidemic growth is not the same as that required to estimate final epidemic size. A seven-day forecast may become empirically evaluable before a 21- or 28-day forecast. Geographic units may become rankable for operational prioritisation before their absolute probabilities of infection or first confirmation can be calibrated. Aggregate deaths and cases may reveal geographic heterogeneity in reported mortality while remaining unable to identify patient-level fatality. Competing generative histories may be compared conditionally even where the available evidence cannot discriminate decisively among the leading mechanisms.

The problem is therefore not simply to determine when an outbreak dataset becomes “mature”. The more operational question is: given the surveillance system, the epidemic information accumulated so far, and a specific decision, what can the available data responsibly support at this point in time?

We define estimand-specific analytical readiness, Rθ,v, as the degree to which the decision, estimand, data vintage, observation process, epidemic information, analytical method and validation evidence jointly support the intended inference. Readiness is therefore a property of an analytical task rather than of a dataset alone: the same information state may support one inference while leaving another model-dependent, scenario-bound or non-identifiable.

Immaturity need not imply that modelling should stop. Instead, it may change the appropriate analytical product. An analyst may need to estimate a reported rather than biological outcome, shorten a forecasting horizon, preserve uncertainty about reporting regimes, provide scenarios instead of a single estimate, rank locations without assigning calibrated absolute probabilities, or state explicitly that the desired quantity cannot be identified from the observations currently available. An apparently precise estimate is not necessarily preferable to a well-supported bound or an explicit conclusion of non-identifiability.

This principle is particularly important during emerging outbreaks, when waiting for complete information is operationally unrealistic but presenting structurally unsupported precision can itself distort response decisions. The appropriate objective is therefore neither to maximise model complexity nor to impose a universal threshold before analysis begins. It is to match the estimand, evidence, model, validation standard and interpretation to the information that is actually available.

In this study, we develop and operationalise an estimand-first, observation-aware framework for analytical readiness in real-time outbreak analysis. Its central claim is deliberately limited: analytical readiness is estimand-specific and must be judged from the observation process, epidemic-process informativeness, method and decision-matched validation, rather than inferred from dataset size or apparent numerical stability. The empirical analyses are structured demonstrations of distinct implications of that claim; they are not presented as universal proof that one fixed indicator set or threshold applies across pathogens and surveillance systems.

Four analytical task families provide the demonstrations: reported-case trend estimation, short-term reported-burden forecasting, geographic prioritisation, and interpretation of reported mortality with latent-fatality identifiability. The unifying contribution is the discipline linking decision, estimand, observation process, S, E, method, validation and permissible interpretation—not a single epidemic “supermodel”.

The 2026 Bundibugyo virus disease outbreak in the Democratic Republic of the Congo provided part of the practical motivation for this work, because different analytical questions arising from the same evolving surveillance system reached different levels of support at different times. It is not, however, the principal object of the present study. The framework is designed and evaluated as a general methodological approach to real-time outbreak analysis under imperfect observation, with the current outbreak used only as a limited contemporary stress-test and worked illustration.

### Conceptual framework

#### Purpose and scope

The framework asks what inference an evolving outbreak surveillance record can responsibly support at a particular data vintage. It does not judge a dataset globally as “good” or “bad” and does not prescribe a preferred model. Instead, it links the operational decision and estimand to the observation process, epidemic signal, method and validation evidence.

The central premise is that analytical validity is conditional on the estimand. The same data may support one task but not another, and a model may fit or converge while the requested quantity remains weakly identified.

Three concepts organise the framework: surveillance maturity (S), describing the reliability and characterisation of the observation system; epidemic-process informativeness (E), describing whether enough relevant epidemic signal has accumulated; and analytical readiness (Rθ,v), describing whether S, E, method and validation jointly support the inference for estimand θ at vintage v.

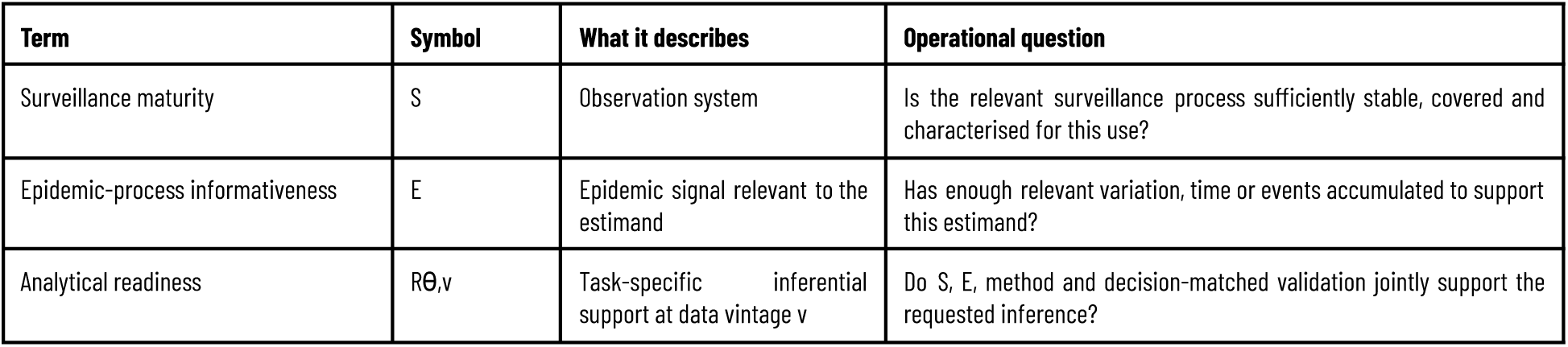
Core definitions used throughout the paper.

#### The estimand-first principle

An estimand is the quantity that an analysis is intended to estimate. Modern statistical guidance has emphasised the importance of defining the scientific question and corresponding estimand before choosing an estimator or analytical procedure [10]. Although formal estimand frameworks have been developed most explicitly for clinical trials, the underlying discipline is directly relevant to outbreak analysis: the target quantity must be specified before deciding what the observed data can be used to estimate.

We extend this principle to real-time outbreak surveillance. An operational question such as “Where will the outbreak spread next?” is not itself a sufficiently precise estimand. Depending on the available data, it could refer to the probability that infection has already been introduced into a location; the probability of biological introduction during a future period; the probability that local transmission becomes established; the probability that the first laboratory-confirmed case will be reported; or the expected rank of a location among those likely to report a case next. These quantities are not interchangeable. A surveillance dataset may support the last two while containing little information about the first three.

Likewise, “What is the fatality rate?” may refer to infection-fatality risk, clinical case-fatality risk among symptomatic cases, fatality among laboratory-confirmed patients with resolved outcomes, or the contemporaneously reported ratio of cumulative deaths to cumulative confirmed cases. Aggregate situation reports may provide the final quantity directly while leaving the others partially or completely unidentified.

We therefore represent the analytical sequence as:

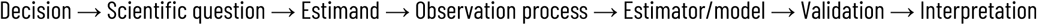

The order is substantive. Selecting a model before defining the estimand risks allowing the structure of the available dataset—or the capabilities of the preferred statistical method—to determine retrospectively what question is claimed to have been answered.

#### Latent epidemic process and surveillance observation process

Let Xi denote the latent epidemic state at time t. Where spatial structure is relevant, let Xz,t denote the state in geographic unit z. Depending on the analytical question, X may include infections, infectious individuals, transmission links, clinical outcomes, deaths, recoveries, geographic introductions or primary spillover events.

These latent quantities are only partially observed. What becomes available to analysts is a surveillance record Y(v), where v denotes the data vintage: the version of the surveillance record that existed at a specified analytical date.

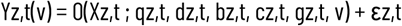

Here q represents detection, ascertainment and testing; d delays between epidemiological events and observation; b laboratory or reporting backlogs and administrative batching; c classification and reclassification; g geographic coverage and attribution; v the information available in the relevant historical data vintage; and ε residual observation error.

This formulation is deliberately general. It is not intended as a single generative statistical model that must be fitted in every analysis. Rather, it requires the analyst to identify which components of the observation process are likely to separate the available surveillance record from the latent quantity of interest.

Public-health surveillance guidance has long recognised attributes such as timeliness, sensitivity, representativeness, data quality and stability as characteristics of the surveillance system rather than properties of the disease itself [3]. The distinction made here is consequently not that surveillance systems distort an otherwise directly observable epidemic—a longstanding concern—but that the consequences of that observation process must be linked explicitly to the particular estimand being requested.

For example, a sudden increase in reported cases could be compatible with an increase in latent transmission, expanded testing, backlog clearance, retrospective case confirmation, or some combination of these mechanisms. Whether those possibilities matter analytically depends on the estimand. They may substantially bias an estimate of instantaneous epidemic growth while having considerably less effect on a longer-term cumulative-burden description.

#### Surveillance maturity (S)

Surveillance maturity (S) is the condition in which the relevant surveillance capacity and processes generate a sufficiently reliable and approximately stable-delay observation regime for the analytical use under consideration. It is a property of the observation system, not a pattern inferred solely from the epidemic curve.

Stable counts cannot by themselves establish S: smooth series can also result from lost access, reduced case finding or a stalled backlog. Capacity- and process-side evidence such as laboratory throughput, reporting participation, delay, backlog, revisions and geographic coverage is therefore used where available.

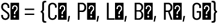

Here C represents surveillance capacity, P operational process performance, L reporting-delay characteristics, B backlog behaviour, R revision burden and G geographic or population coverage.

S may vary by time, geography, outcome and reporting channel and can regress after insecurity, laboratory interruption, case-definition change or loss of coverage. The framework therefore never classifies an entire country or outbreak permanently as mature or immature.

#### Epidemic-process informativeness (E)

S addresses whether the observation mechanism is sufficiently reliable; it cannot determine whether the epidemic has generated enough information for the intended analysis.

Epidemic-process informativeness (E) is the amount of estimand-relevant signal generated by the epidemic, including elapsed time, event counts, trajectory variation, affected geographies, resolved outcomes and matured historical predictions.

The relevant components of E are task-specific: long-horizon forecasting may require matured forecast origins and regime stability, while fatality inference requires resolved outcomes and information that separates ascertainment from outcome risk.

This paper isolates E analytically from S so that observation-system adequacy and epidemic information are not conflated. A fully functioning surveillance system cannot supply curvature, resolved outcomes or future forecast errors that have not yet occurred.

#### A two-dimensional readiness space

Surveillance maturity and epidemic-process informativeness define two conceptually independent dimensions.

**Figure 1.**
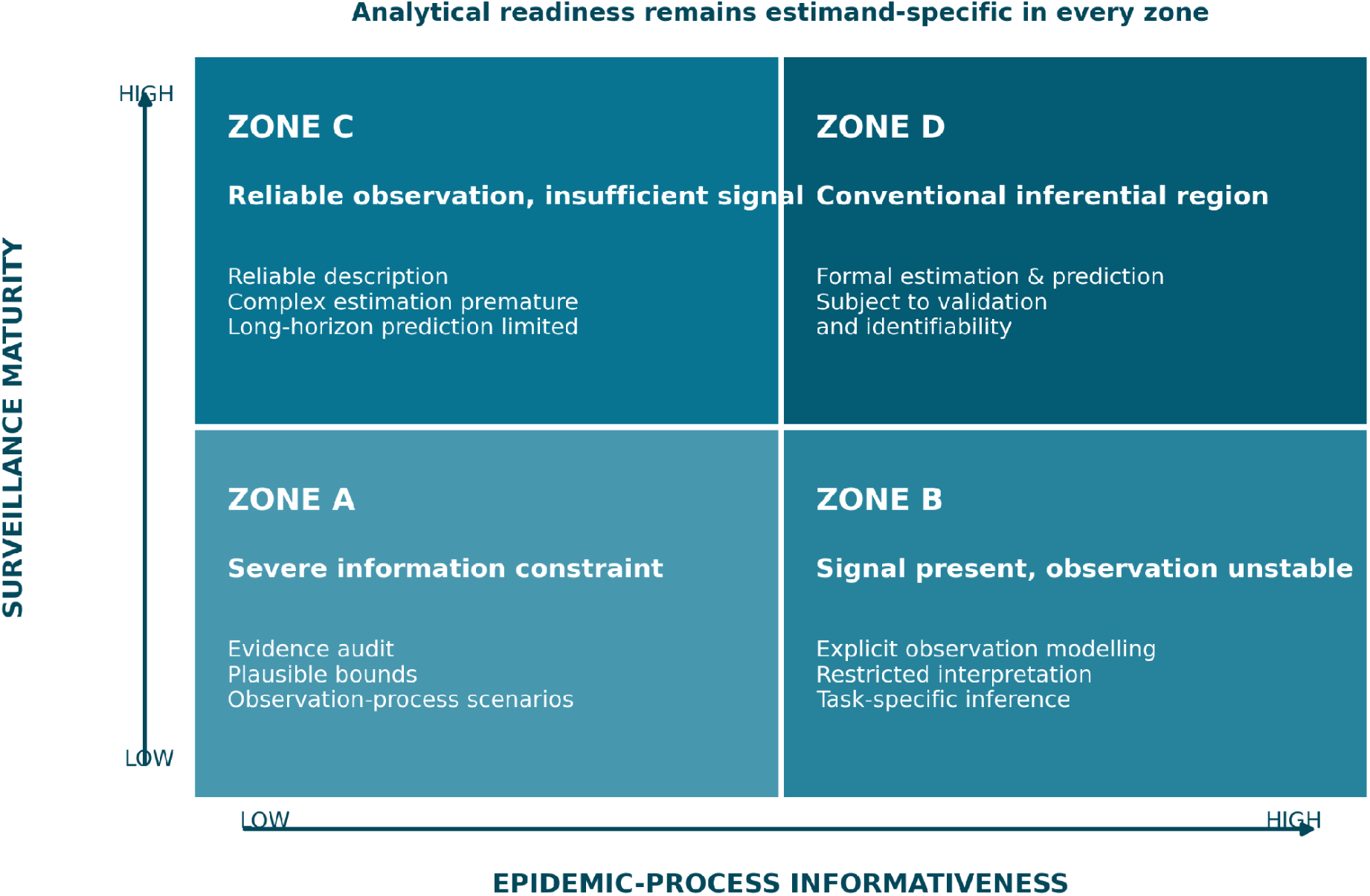
Conceptual analytical-readiness space. The two dimensions are surveillance maturity (S) and epidemic-process informativeness (E). Quadrants are conceptual regions rather than discrete biological states; estimand-specific readiness still depends on method, validation and identification.

Outbreaks may move gradually between them and may move backwards along the surveillance dimension.

More importantly, location within the two-dimensional space does not itself determine whether a particular analysis is permissible. A task-specific third element is needed: analytical readiness. Thus, an outbreak in Zone B may still permit a useful short-term forecast if the estimand is explicitly future reported cases and the model represents the surveillance change. The same data may not support inference about underlying infection incidence. Similarly, Zone C may provide high-quality case counts but remain unable to identify final epidemic size because insufficient curvature has yet developed.

#### Estimand-specific analytical readiness (Rθ,v)

Analytical readiness Rθ,v is the support available for a specified inference about estimand θ at data vintage v.

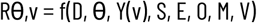

Here D is the operational decision, θ the estimand, Y(v) the available data vintage, S surveillance maturity, E epidemic-process informativeness, O the material observation-process assumptions, M the method and V the validation evidence.

Different estimands may achieve readiness at different times during the same outbreak. Early growth may become estimable before saturation. A one-week forecast may be empirically defensible before a three-week forecast. Geographic rankings may become actionable before absolute hazards are calibrated. Reported mortality heterogeneity may be measurable while clinical fatality remains unidentified.

Analytical readiness can also decline. A surveillance disruption, geographic expansion into poorly observed areas or a change in case definition can invalidate a previously adequate observation model. Readiness is therefore evaluated repeatedly as new data vintages become available rather than declared once for the duration of an outbreak.

#### Identifiability as a separate requirement

A model may fit the available observations without the quantity of interest being identifiable. We therefore treat identifiability as distinct from convergence, goodness-of-fit and predictive adequacy.

The methodological literature commonly distinguishes structural identifiability, where different parameter combinations can generate indistinguishable observations even with ideal data, from practical identifiability, where the model may be structurally distinguishable in principle but the available data are too sparse, noisy or weakly informative to identify its parameters reliably [11].

We adapt this distinction to outbreak surveillance. An estimand may be structurally non-identifiable from the available data type. For example, two aggregate totals—reported confirmed cases and reported confirmed deaths—cannot independently identify true infection ascertainment, death ascertainment, unresolved outcomes and patient-level fatality without additional information or assumptions. Alternatively, an estimand may be practically non-identifiable at the current outbreak stage. A saturation parameter may eventually become estimable from a complete epidemic trajectory but remain effectively unconstrained during early exponential growth.

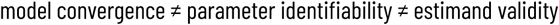

Successful numerical convergence is a property of the fitting procedure. Identifiability concerns whether the observations distinguish the relevant parameters or mechanisms. Estimand validity additionally requires that those parameters correspond to the scientific quantity being claimed.

#### Analytical output classes

Rθ,v is expressed through six non-hierarchical inferential-support classes: A, empirically supported estimation; B, estimation that remains materially model-dependent; C, scenario-bound inference; D, validated ranking without calibrated absolute probabilities; E, a directly observable surveillance quantity without justified biological interpretation; and F, non-identifiability from the available evidence. The classes describe different permissible outputs rather than grades of dataset quality; their operational evidence rules are specified in Section 4.3 and Supporting Information S2.

#### Decision-matched validation

Validation should address the intended use rather than generic fit. Forecasting requires time-respecting predictive evaluation; prioritisation requires decision-matched ranking metrics such as Top-k capture; latent clinical quantities require identification and ascertainment sensitivity; and competing-mechanism analyses, where used, require model-recovery evidence.

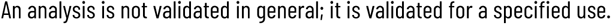

#### Data vintage as part of the observation process

Real-time outbreak datasets evolve retrospectively. Cases may be added to earlier dates, removed as duplicates, reclassified, geographically reassigned or corrected after laboratory reconciliation. Consequently, the dataset available today describing day t may not be the dataset that was available on day t.

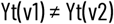

Outcome leakage occurs when future outcomes are used directly or indirectly to fit a prediction made at an earlier analytical origin. Data-vintage leakage occurs when a retrospective analysis uses later corrections to the historical input data and treats those corrected values as though they had been available contemporaneously.

A rolling-origin evaluation may avoid the first while retaining the second. The distinction between true prospective evaluation and retrospective or pseudo-prospective reconstruction should therefore depend not merely on whether later target outcomes were withheld, but also on whether historically appropriate data vintages were available.

This concern is not merely theoretical: recent re-analysis has shown that using consolidated surveillance data in retrospective epidemic forecasts can create information leakage when data revisions are ignored [12].

#### The analytical-readiness workflow

Figure 2 is the organising workflow for the paper. The analyst begins with the operational decision and scientific question, defines the estimand, identifies what is actually observed and characterises the observation process, then evaluates S and E, chooses a defensible method, and applies validation matched to the intended decision. Rθ,v is the resulting judgement about the strongest inference the evidence can support.

**Figure 2.**
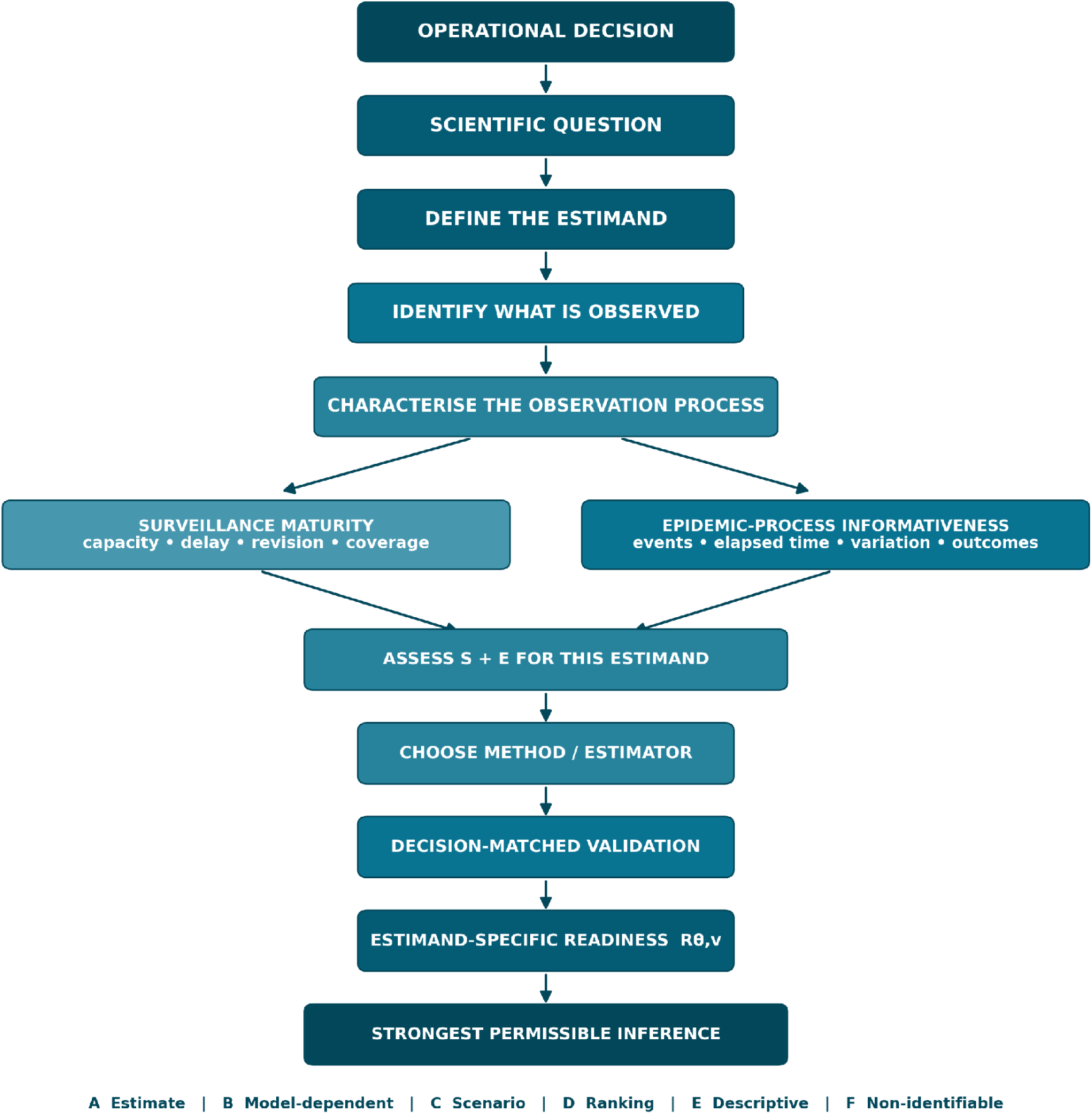
Estimand-first analytical-readiness workflow. The workflow links the operational decision and scientific question to the estimand, observed data, observation process, S and E, method, decision-matched validation, Rθ,v and the strongest permissible inference.

#### What the framework is—and is not

The framework is deliberately a discipline for matching evidence to claims, not a new universal scoring instrument.

| The framework is | The framework is not |
| --- | --- |
| A way to align a decision, estimand, data vintage, observation process and validation standard. | A single statistical model or mandatory observation model. |
| A task-specific judgement that can change across estimands and time. | A global label that a dataset, outbreak or country is “ready”. |
| A rule for weakening or changing the analytical product when evidence is limited. | A requirement to stop analysing until surveillance is perfect. |
| A framework in which non-identifiability can be the correct result. | A scalar maturity score or an ordinal league table of data quality. |

### Study aim and evaluative propositions

#### Study aim

The study aims to operationalise analytical readiness as an estimand- and vintage-specific judgement and to test whether that judgement behaves coherently across distinct outbreak-analytic tasks. The empirical datasets are structured demonstrations: each tests a specific implication of the framework rather than serving as universal validation of the entire architecture.

#### Primary research question

At a given outbreak information state, what inference can the available surveillance record responsibly support for a specified estimand and decision, once the observation process, epidemic-process informativeness and relevant validation evidence are taken into account?

#### Evaluative propositions

P1 — Estimand specificity and temporal dynamics. Analytical readiness is not dataset-wide: different estimands can occupy different readiness states at the same data vintage, and Rθ,v can increase or decrease as S, E and the observation regime change.

P2 — Decision-matched validation. Analytical support depends on the task, horizon, comparator and validation evidence; additional historical data or greater model complexity do not by themselves establish readiness.

P3 — Precision is not identification. Numerical convergence, apparent stability or narrow uncertainty around an observed surveillance statistic do not establish that the desired latent estimand is identified.

P4 — Evidence-adaptive analytical products. Imperfect surveillance should change the permissible analytical product and claim strength—empirically supported or model-dependent estimation, scenarios, ranking, description or non-identifiability—rather than force a binary ready/not-ready decision.

#### Comparative strategy

The DRC surveillance-shock analyses test P1 by comparing forecast and trend behaviour across a documented deterioration and recovery in surveillance; the geographic prioritisation analysis tests P2 and P4 by comparing probability scoring with the Top-k allocation decision; the independent Western Area analysis tests P2 by examining horizon dependence and transparent baselines in a second outbreak; and the mortality experiment tests P3 and P4 where latent truth is not empirically observable.

No single module is expected to prove the framework as a whole. Support is instead sought through coherence across targeted implications: readiness should vary by estimand when the evidence differs; decision-matched validation should alter conclusions when the operational task changes; identification diagnostics should prevent precise observed quantities from being over-interpreted; and restricted evidence should lead to restricted claims rather than unsupported point estimates.

#### Criteria for interpretation

The framework is supported to the extent that contemporaneous readiness judgements distinguish analyses that later remain stable or perform adequately from those that become unstable, miscalibrated or non-discriminatory, and that the same data can correctly yield different permissible outputs for different estimands. Results are treated as demonstrations of these propositions, not as proof of universal thresholds or indicator weights.

#### Role of the 2026 Bundibugyo virus disease outbreak

The 2026 Bundibugyo virus disease outbreak remains a contemporary motivation and worked illustration rather than the principal validation dataset. It was not used to tune thresholds or define universal readiness rules.

## Methods

### Study design and validation map

We conducted a longitudinal, multi-outbreak methodological study combining repeated analysis of successive real-world surveillance states with a targeted simulation experiment where the latent quantity could not be observed directly. The principal unit was U=(o,T,θ): outbreak o, analytical origin T and estimand θ. At each origin, the question was not whether a dataset was globally “ready”, but what inference was supportable for θ given the data vintage, observation process, epidemic-process information, method and validation available at that time.

Four task families were evaluated: reported-case trend estimation, short-term reported-burden forecasting, geographic prioritisation and interpretation of reported mortality with latent-fatality identifiability. They were deliberately not pooled into a single accuracy score because each task has a different estimand and validation standard. Table 1 maps the four demonstrations to the proposition each was intended to test. The full predefined estimand–evidence–validation matrices and implementation rules are provided in Supporting Information S2.

**Table 1.** Validation map for the four empirical demonstrations.

| Demonstration | Operational estimand | Data / origin structure | Primary validation or diagnostic | Framework proposition |
| --- | --- | --- | --- | --- |
| DRC trend stability | Four-week multiplicative trend in reported confirmed cases | Successive origins across the Nov-Dec 2019 surveillance shock | Window sensitivity, overdispersion and variance-adjusted uncertainty | P1, P3 |
| Reported-burden forecasting | Future reported burden at predefined horizon: 7-day cumulative DRC cases; 1-4-week Western Area incidence | DRC shock origins plus independent Western Area rolling origins | Absolute error, horizon dependence, transparent baseline and published calibration evidence | P1, P2 |
| Geographic prioritisation | Rank of six surveyed health zones for $\geq 2$ reported cases during Dec 2019 | Archived DRC forecasts issued 15 Nov-1 Dec | Top-k capture versus cumulative-burden comparator; Brier score secondary | P2, P4 |
| Mortality identifiability | Reported rCFR versus latent fatality p | Targeted structural and stochastic experiment | Equivalence mechanisms and precision-versus-identification analysis | P3, P4 |

### Data sources, vintages and observation-process evidence

The 2018–2020 Ebola virus disease outbreak in North Kivu and Ituri, Democratic Republic of the Congo (DRC), provided the principal historical observation-process stress test because it combined a long health-zone surveillance series with contemporaneous documentation of laboratory, alert, contact-follow-up and access conditions [13]. The 2014–2015 Western Area epidemic in Sierra Leone provided an independent forecasting dataset with a preserved 27-week incidence series and published real-time forecast evaluation [8]. The 2026 Bundibugyo virus disease record motivated the framework and supplied limited worked illustrations, but it was not used to tune readiness rules or as the principal validation dataset.

Historical information was treated as part of the observation process rather than as a frictionless record of the latent epidemic. Priority was given to contemporaneous situation reports and operational updates; digitised repositories derived from those materials were cross-checked where discrepancies were relevant to the estimand. Missing reports were not interpreted as zero events, and disagreements in contemporaneous values or geographic attribution were retained as provenance uncertainty rather than silently harmonised. Where later-corrected data could not be separated from the record available at the analytical origin, the analysis was described as pseudo-prospective rather than vintage-faithful.

Surveillance maturity S was assessed as a profile of observation-system evidence rather than a scalar index. Relevant evidence included reporting continuity, timeliness, backlog or batching, surveillance activity, revision burden, geographic coverage and cross-channel consistency. Epidemic-process informativeness E was assessed from estimand-relevant information such as elapsed time, event counts, variation, affected geographies, matured forecast origins and resolved outcomes. The detailed indicator catalogue, vintage taxonomy, missingness rules and transition logic are reported in Supporting Information S1; the source hierarchy and DRC provenance audit are in S3.

### Readiness assessment, inferential-support classes and analytical specification

For each outbreak, origin and estimand, analytical readiness Rθ,v was judged from the decision, the estimand, the available data vintage, S, E, material observation-process assumptions, the analytical method and task-matched validation. The judgement was recorded as the strongest inferential-support class justified by the complete evidence profile. Classes A–F are non-hierarchical: they specify the kind of result that may be reported, not a numeric quality score.

**Table 2.** Non-hierarchical inferential-support classes.

| Class | Typical evidence profile | Permissible analytical output | Claims that remain unsupported |
| --- | --- | --- | --- |
| A – Empirically supported | Estimand aligned with observation process; no material unresolved discontinuity in the relevant window; robust across defensible specifications; decision-matched empirical validation adequate | Numerical estimate or predictive distribution with uncertainty appropriate to the use | Claims beyond the validated estimand, horizon or population; assumption-free certainty |
| B – Materially model-dependent | Meaningful signal exists, but conclusions vary with plausible priors, windows, delay models, observation assumptions or limited external validation | Numerical estimate with model dependence visible in the primary result and sensitivity analysis | Presentation as uniquely identified, fully robust or generally validated |
| C – Scenario-bound | Observations constrain the estimand only conditional on substantively different assumptions that cannot be defensibly weighted | Scenario-specific estimates or bounded range with assumptions shown explicitly | A single preferred point estimate obtained by averaging unsupported scenarios |
| D – Rankable, not probability-calibrated | Relative ordering repeatedly improves the decision-matched ranking metric versus a relevant comparator, while absolute probabilities or timing remain unvalidated | Priority ranking or Top-k allocation product | Calibrated absolute event probabilities, exact timing or causal mechanism |
| E – Descriptively observable | Surveillance statistic is directly observed, possibly with high precision, but the corresponding latent biological quantity is not identified | Reported surveillance quantity with explicit descriptive interpretation | Biological incidence, fatality, introduction date or other latent interpretation not identified by the observation process |
| F – Non-identifiable | Materially different latent states/mechanisms remain observationally equivalent or recovery fails under defensible specifications | Explicit non-identifiability result plus the additional information required for identification | A preferred latent point estimate or mechanism presented as data-resolved |

Classification rules were fixed before cross-module synthesis and were not optimised against the observed comparative results. Quantitative indicators were retained continuously where possible; documented backlogs, missing historical vintages and other categorical observation-process features were recorded explicitly rather than converted into arbitrary weights. Readiness could increase, remain unchanged or decline across successive origins. The full evidence rules, transition definitions and worked decision matrices are supplied in Supporting Information S1–S2.

The study was not prospectively preregistered. For the historical stress test, analytical origins, target estimand, persistence comparator and observation-aware product rule were specified before the corresponding forecast-error comparison was interpreted; class definitions and validation logic were fixed before cross-module synthesis.

#### Worked classification example: O3 reported-case trend

At the O3 recovery origin, the estimand was the multiplicative four-week trend in reported confirmed cases. Surveillance activity had substantially recovered, but the recent observation regime had changed and the fitting window contained the large 4–10 December reporting/transmission episode. The Poisson fit converged and produced a weekly multiplier of 1.29, yet Pearson dispersion was 7.65 and variance adjustment widened the 95% interval to 0.65–2.55. The trend therefore failed the robustness requirement for Class A but retained estimand-relevant information: it was classified as Class B and reported as materially model-dependent rather than as a resolved increase in latent transmission.

### DRC surveillance-shock forecast and trend experiment

A quasi-experimental window was specified before the corresponding forecast-error comparison around the November–December 2019 security disruption in the North Kivu/Ituri Ebola response. The window was selected from contemporaneous WHO process evidence rather than from forecast error. WHO documented a sharp reduction in alerts and contact follow-up after violence and response suspension in late November, followed by partial recovery in early December and a return toward previous operational levels by mid-December [15–18]. Four analytical origins were fixed: 19 November (O0, pre-shock), 26 November (O1, acute shock), 3 December (O2, early recovery) and 17 December (O3, recovery).

#### Seven-day reported-burden forecast

The forecast estimand was the cumulative number of reported confirmed cases seven days after origin T, equivalently the seven-day change in the reported cumulative series. This was explicitly an observation-level target, not infection or onset-date incidence. A fixed persistence comparator projected the most recent seven-day reported increment forward:

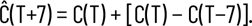

Absolute error against the subsequently reported cumulative total was the primary metric. The same comparator was evaluated in the Beni–Mabalako–Oicha corridor, a four-zone active core adding Mandima, and an aggregate across all listed health zones. The observation-aware product rule did not “correct” the epidemic curve: it allowed numerical forecasting before the disruption, withheld a calibrated point forecast during acute disruption, continued to withhold during early recovery until the observation regime demonstrated renewed stability, and allowed numerical forecasting to resume at O3 with explicit model-dependence. Product choices were based on contemporaneous process evidence rather than subsequent performance.

#### Rolling reported-case trend

At the same four origins, a second product estimated the local trend in reported confirmed cases using the four most recent completed WHO reporting weeks. A Poisson log-linear model was fitted,

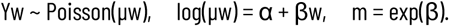

The principal estimand m was the multiplicative weekly trend in the reported case stream, not a reproduction number or latent infection-growth rate. Pearson dispersion φ was calculated from each four-point fit. Standard errors were not reduced when φ<1 and were multiplied by sqrt(φ) when φ>1. A three-week fitting window was specified as a sensitivity analysis before cross-module synthesis. This paired design tested whether forecast readiness and trend-estimation readiness moved synchronously through the same observation-process disturbance. Detailed origin construction, source reconciliation and calculations are in Supporting Information S3–S4.

### Archived geographic-prioritisation stress test

A secondary spatial analysis used committed December 2019 health-zone forecasts from the Ebola Expert Elicitation repository [14]. The archived target was not biological introduction or first confirmation. It was whether each of six surveyed health zones—Beni, Goma, Kalunguta, Mabalako, Mambasa and Mandima—accumulated at least two reported cases during December 2019. Four zones subsequently met that target.

Forecasts issued on 15 November, 20 November, 22 November and the nominal 1 December date were evaluated for a plain EpiCastR specification and an adjacency-interaction specification. A transparent comparator ranked the same six zones by cumulative cases already reported at the forecast date. The primary operational metrics were the number of realised-positive zones captured in the Top 3 and Top 4 priorities; mean Brier score was retained as a secondary probability metric. Because the stress test contained six zones and one target month, probability scores were interpreted descriptively. A code audit identified a nominal-date indexing discrepancy in the archived adjacency scoring workflow; the 1 December adjacency probabilities were therefore read directly from the committed raw forecast file. Full probabilities, rankings and the provenance correction are documented in Supporting Information S5.

### External forecast validation in Western Area, Sierra Leone

External forecasting evidence used the preserved data and analysis package accompanying Funk et al. [8]. The Western Area series contained 27 weekly reported-incidence observations beginning with the week ending 10 August 2014. Retrospective forecast assessment began at week 3, consistent with the published analysis. The source reconstruction also preserved an observation-process trade-off: recent observations were taken from timely Ministry situation reports, whereas older observations came from the continuously cleaned but delayed WHO patient database.

A transparent rolling-origin persistence forecast set future reported weekly incidence equal to incidence at origin T. Forecasts were evaluated at one- to four-week horizons using absolute error and symmetric absolute percentage error. For each origin, HT was the number of completed weekly observations and the descriptive history-to-horizon ratio was MT,h=HT/h. M was examined continuously and in predefined bins (<1, 1–<2, 2–<4, 4–<8, >=8); no bin was treated as a readiness threshold. Reconstructed persistence errors were then compared with the published calibration assessment and median absolute errors for the semi-mechanistic and null models. The aim was to test horizon dependence, the value of transparent baselines and whether M>=1 could be treated as sufficient for readiness. Detailed rolling origins and calculations are in Supporting Information S6.

### Mortality-identifiability experiment

For mortality, the directly observed quantity was the reported crude case-fatality ratio rCFR=Dreport/Creport. The study did not assume that this ratio represented patient-level fatality. To isolate structural identification, let N denote true infections or relevant clinical cases, p latent fatality, qC case ascertainment, qD death ascertainment and u the fraction of eventual fatal outcomes observable by the analytical origin. Under the simplified observation system,

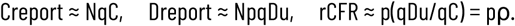

The structural illustration fixed Creport=1,000 and Dreport=400 (rCFR=0.40), evaluated qC and qD on a 0.25–1.00 grid and varied u from 1.0 to 0.4. Five explicit latent mechanisms were constructed to have the same expected reported counts while true fatality ranged from 0.10 to 0.80. For each mechanism, 5,000 stochastic replicates (seed 260810) sampled reported cases from Binomial(N,qC), true deaths from Binomial(N,p), and observed deaths from Binomial(true deaths,qDu).

A second experiment separated sampling precision from identification. At a fixed reported ratio of 0.40, reported-case denominators ranged from 100 to 25,000. A Wilson interval, used only to represent sampling precision of the reported ratio, was compared with the structural fatality range under the predefined effective relative-ascertainment bound ρ=qDu/qC in [0.5,2], implying p in [0.20,0.80]. The diagnostic was whether the reported-ratio interval contracted while the structural identification interval remained unchanged. The full equivalence grid, mechanism definitions and simulation outputs are in Supporting Information S7.

### Statistical interpretation, sensitivity, reproducibility and ethics

The study was methodological and comparative rather than organised around a single null-hypothesis test. Repeated analytical origins from the same outbreak were not treated as independent replicates. Each module therefore used task-matched uncertainty and sensitivity analyses: three aggregation levels for the DRC forecast, alternative trend windows and dispersion adjustment, plain versus adjacency spatial specifications and Top-3 versus Top-4 decisions, one- to four-week Sierra Leone horizons, and explicit mortality ascertainment/outcome-maturity scenarios. Disagreement between metrics was retained rather than resolved through post hoc selection.

An accompanying reproducibility archive preserves compact analysis-ready inputs, source lineage, deterministic analysis rules, Python and R implementations, expected outputs, figure-generation scripts and file checksums. The empirical analyses used aggregated, de-identified surveillance information and public historical archives; no identifiable individual-level information was required, and the targeted simulation used no human-subject data. Extended reproducibility, implementation and audit specifications are in Supporting Information S8.

OpenAI ChatGPT and Claude (Anthropic, Claude Sonnet 5) were used as assistive tools during manuscript drafting and editing, document formatting, citation verification, and code-generation support. JGLV reviewed and verified the analytical code, numerical outputs, references, interpretations and final text. The authors retain full responsibility for the work; neither system was assigned authorship.

## Results

### Historical data provenance and observation-process evidence

The historical DRC data architecture was adequate for an empirical-first evaluation. The Epiforecasts archive preserves a daily health-zone surveillance series derived from Ministry of Health/WHO/HDX reporting and earlier extracts of the same data lineage. The archived series retains corrective changes in cumulative counts, including occasional decreases in previously reported confirmed totals. These were treated as evidence of revision or reclassification in the observation process rather than as negative incidence.

Contemporaneous WHO reports additionally provided process-side evidence independent of the epidemic curve. On 28 November 2019, WHO reported that violence and civil unrest had restricted response access; daily alerts in Beni had fallen from approximately 400 to 120–150, the overall proportion of contacts under surveillance had fallen as low as 59% on 25 November, and only 15% of contacts were under surveillance in Oicha on 26 November [15]. One week later, 96% of 3,346 reported alerts were investigated within 24 hours, but the average proportion of contacts under surveillance remained 70%, including 42% in Oicha; WHO described the indicators as improving slowly but continuing to fluctuate [16]. By 10–17 December, WHO reported that alert volume and contact-surveillance performance had returned toward levels observed before the security events, although Mabalako still had the lowest contact follow-up, at 82% on 17 December [17,18].

This produced a natural sequence in which surveillance performance moved sharply downward and then upward while chronological epidemic history continued to accumulate: pre-shock observation, acute disruption, early recovery, and operational recovery. The sequence was therefore used as the first empirical test of whether analytical readiness can change independently of simple elapsed outbreak time.

### Paired seven-day forecast-readiness experiment

At the three-zone corridor level, the seven-day persistence comparator produced mean absolute errors of 0.67 cases at the pre-shock origin, 0.67 during acute disruption, 7.33 during early recovery, and 1.67 after recovery. The largest error therefore did not occur at the point of greatest documented surveillance disruption. It occurred one week later, when process indicators were improving but the observation regime had not yet demonstrated renewed stability.

The same pattern persisted under broader aggregation. At the all-health-zone level, reported confirmed totals progressed from 3,173 on 12 November to 3,180 on 19 November, 3,186 on 26 November, 3,195 on 3 December, 3,222 on 10 December, 3,233 on 17 December, and 3,248 on 24 December. The totals through 17 December matched contemporaneous WHO confirmed totals on the corresponding reporting dates [15–20].

At the 3 December early-recovery origin, the preceding all-health-zone seven-day increment was 9 cases (3,186 to 3,195). Persistence therefore predicted 3,204 confirmed cases on 10 December. The subsequently reported total was 3,222, corresponding to an increment of 27 and an absolute forecast error of 18 cases. In contrast, all-health-zone absolute errors were 1 case at the pre-shock origin, 3 during the acute shock, and 4 after recovery.

**Table 3.** Seven-day persistence forecast error across surveillance phases.

| Origin | Surveillance phase | 3-zone corridor | 4-zone active core | All-HZ aggregate |
| --- | --- | --- | --- | --- |
| 00 | Pre-shock | 2 | 1 | 1 |
| 01 | Acute shock | 2 | 3 | 3 |
| 02 | Early recovery | 22 | 18 | 18 |
| 03 | Recovery | 3 | 3 | 4 |

Note. Errors are absolute differences between the fixed seven-day persistence forecast and the subsequently reported cumulative confirmed-case count. The estimand is future reported confirmed burden, not latent infection incidence.

**Figure 3.**
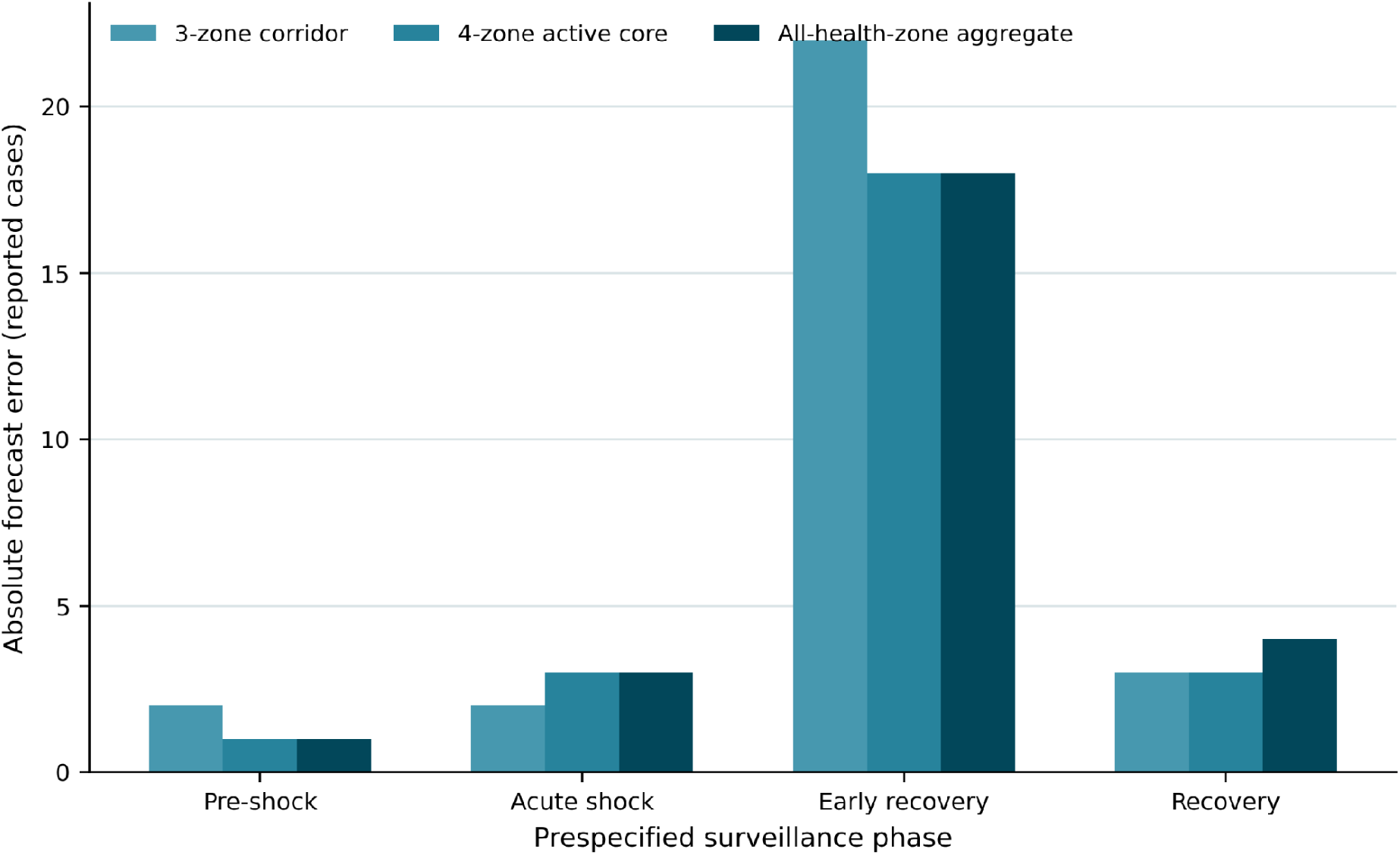
Seven-day persistence forecast error across surveillance phases. Absolute error is shown for three spatial aggregation levels at four specified surveillance origins. The estimand is future reported confirmed burden, not latent infection incidence.

### Concentration of the early-recovery instability

The early-recovery aggregate miss was strongly concentrated but was not purely a single-zone artefact. Between 26 November and 3 December, the all-health-zone reported confirmed total increased by 9 cases; between 3 and 10 December it increased by 27. The resulting increment surprise of +18 cases decomposed arithmetically into +16 in Mabalako, +5 in Beni, +1 in Oicha, −4 in Mandima, and zero net change across the remaining health zones. Mabalako therefore accounted for 16/18 (88.9%) of the net aggregate surprise, while changes in Beni and Oicha also contributed and Mandima partially offset them.

The geographic decomposition also exposed a provenance distinction. WHO reported 27 new confirmed cases from 4–10 December and allocated them as 18 in Mabalako, 6 in Beni, 2 in Mandima and 1 in Oicha [17]. Arithmetic differences in the archived cumulative health-zone series yielded 19, 5, 2 and 1, respectively, while preserving the same national total of 27. The disagreement therefore concerned geographic/event-time attribution rather than overall reported burden. It was retained as an observation-process discrepancy rather than reconciled post hoc.

### Observation-aware product choice and the cost of conservatism

The observation-aware rule would have allowed numerical forecasting at the pre-shock origin, withheld a calibrated point forecast during the acute disruption, continued to withhold a point forecast during early recovery, and allowed numerical forecasting to resume at the recovery origin with an explicit model-dependence caveat. This rule was based solely on contemporaneous surveillance evidence.

Ex post, this rule would have protected against the largest numerical miss: the early-recovery origin, where the all-health-zone naive absolute error was 18 cases. However, it would also have withheld a point forecast at the acute-shock origin where the same naive comparator happened to perform reasonably well, with an all-health-zone absolute error of 3 cases. The first empirical test therefore identifies a genuine trade-off between false readiness and false non-readiness rather than showing that surveillance-based gating mechanically improves numerical accuracy at every origin.

This distinction is central to the proposed framework. A point forecast that proves accurate after the fact does not establish that numerical confidence was justified when the observation system was demonstrably disrupted. Conversely, restricting inference during disruption carries an opportunity cost when an otherwise useful numerical forecast would have performed adequately. Readiness assessment should therefore be judged by whether it reduces unjustified confidence at acceptable decision cost, not by whether every restricted forecast would have been wrong.

### Parameter stability differs from forecast stability

The rolling four-week reported-case trend changed substantially across the same origins. At O0, the estimated weekly multiplier was 0.67 (variance-adjusted 95% CI, 0.51–0.88), consistent with a declining reported case stream. At O1 it was 0.73 (0.53–1.00). At the early-recovery origin O2, the point estimate moved to 1.12 but remained imprecise (0.80–1.56). At O3, after the 27-case week of 4–10 December had entered the fitting window, the point estimate was 1.29, but Pearson dispersion increased to 7.65; variance adjustment widened the interval to 0.65–2.55.

This contrast is important for identifiability. Without the dispersion adjustment, the conventional four-point Poisson fit at O3 yields a 95% interval for the weekly multiplier of approximately 1.01–1.65, which could be read as evidence of increasing incidence. Once the variance visible in the same observations is acknowledged, the data no longer distinguish a sustained increase from decline with useful precision. The model can therefore converge and produce a seemingly decisive slope while the trend estimand remains materially model-dependent, consistent with P3.

**Table 4.** Rolling reported-case trends across surveillance-shock origins.

| Origin | Phase | Four weekly counts | Weekly multiplier | Variance-adjusted 95% CI | Pearson dispersion | Three-week sensitivity |
| --- | --- | --- | --- | --- | --- | --- |
| 00 | Pre-shock | 19, 15, 6, 7 | 0.67 | 0.51–0.88 | 0.69 | 0.64 (0.37–1.11) |
| 01 | Acute shock | 15, 6, 7, 6 | 0.73 | 0.53–1.00 | 0.89 | 1.00 (0.58–1.73) |
| 02 | Early recovery | 6, 7, 6, 9 | 1.12 | 0.80–1.56 | 0.18 | 1.15 (0.69–1.92) |
| 03 | Recovery | 6, 9, 27, 11 | 1.29 | 0.65–2.55 | 7.65 | 1.07 (0.31–3.66) |

*Note. The weekly multiplier is $\exp(\beta)$ from a four-week Poisson log-linear model of WHO-reported confirmed cases. Intervals use the Poisson standard error unless Pearson dispersion exceeds 1, in which case the standard error is inflated by $\sqrt{\phi}$ . Values describe the reported-case stream, not latent transmission.*

**Figure 4.**
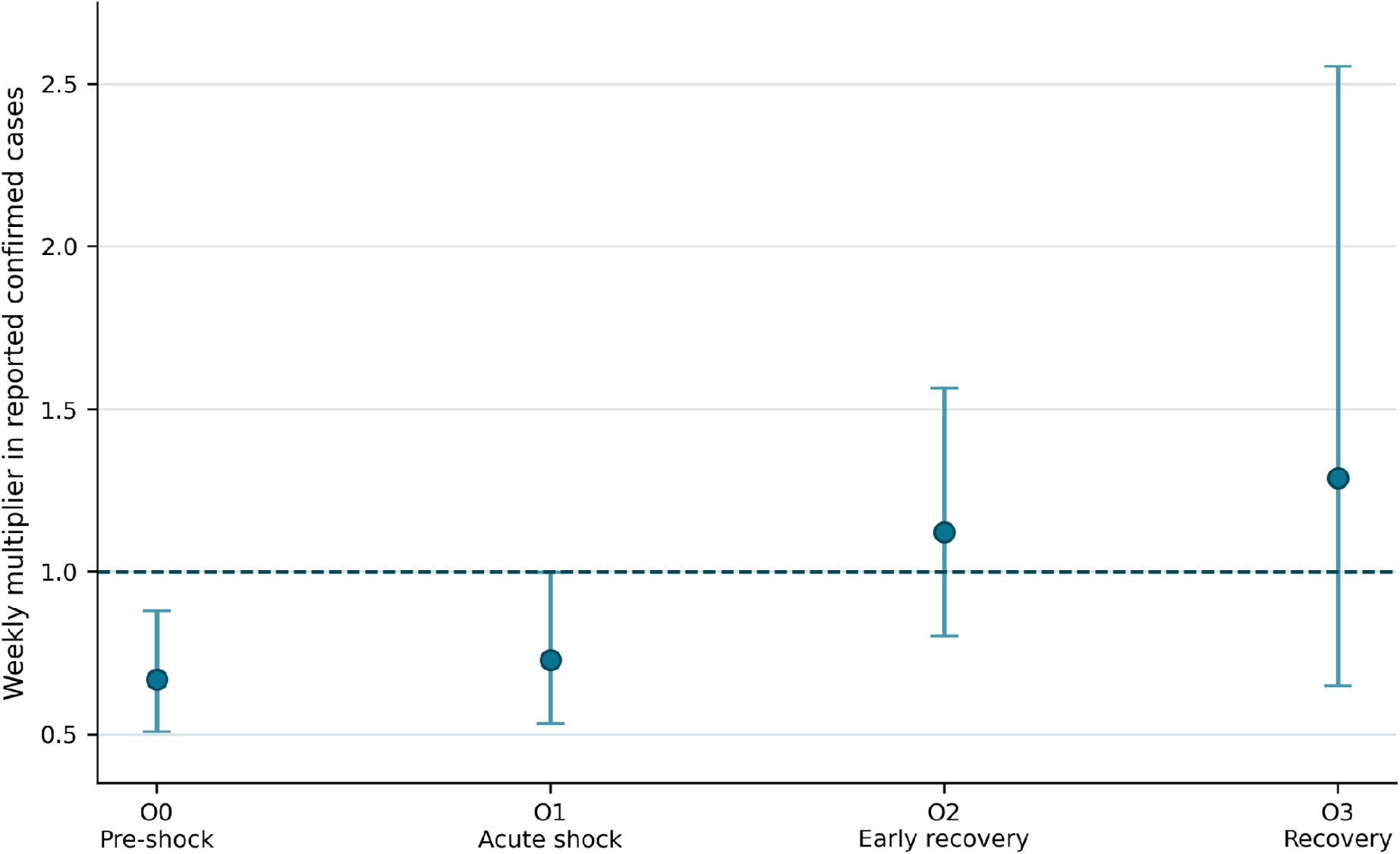
Rolling reported-case trends across surveillance-shock origins. Points are four-week Poisson log-linear weekly multipliers; intervals are variance-adjusted 95% confidence intervals. The dashed reference is a multiplier of 1.0.

The timing of this instability differed from the forecast result. The largest seven-day forecast error occurred at O2, when the rolling trend estimate was still close to flat and its interval spanned both decline and increase. The strongest overdispersion and apparent trend shift emerged at O3, after the large 4–10 December episode entered the estimation window. Forecast readiness and trend-estimation readiness therefore did not move synchronously within the same surveillance sequence, providing direct empirical support for P1.

The parameter shift cannot be attributed to surveillance recovery alone. WHO reported 27 confirmed cases during 4–10 December, all linked to three transmission chains, with 17 infections potentially linked to one individual; the following week, all 11 newly reported cases were linked to the same relapse-associated chain [17,18]. The period therefore contains both documented recovery of surveillance functions and a genuine change in the epidemic process. The experiment is a natural stress test of analytical readiness, not a causal instrument for surveillance quality.

### Rank stability does not by itself establish geographic readiness

Among the six surveyed health zones, Beni, Kalunguta, Mabalako and Mambasa subsequently accumulated at least two reported cases during December; Goma and Mandima did not. The cumulative-burden comparator selected Beni, Mabalako and Mandima as its top three at each analysed origin, thereby capturing two of the four subsequently positive zones. Adding Kalunguta as the fourth priority increased capture to three of four.

The plain spatial model improved on this Top-3 decision only at the 15 November pre-shock origin, when its three highest risks were assigned to Mabalako, Beni and Kalunguta and all three subsequently met the target. At 20 November, 22 November and 1 December, Mandima entered the model Top 3 despite remaining below the realised December threshold, reducing Top-3 capture to two of four. Top-4 capture was three of four under every date and specification. Thus, the risk ordering remained operationally stable in a broad sense, but the pre-shock gain over a simple burden ranking was not sustained through the disruption window.

**Table 5.** Decision-matched performance of geographic prioritisation forecasts.

| Forecast date | Model | Top-3 hits / 4 | Top-4 hits / 4 | Mean Brier | Rank correlation with burden |
| --- | --- | --- | --- | --- | --- |
| 15 Nov | Plain | 3 | 3 | 0.220 | 0.851 |
| 15 Nov | Adjacency | 2 | 3 | 0.199 | 0.943 |
| 20 Nov | Plain | 2 | 3 | 0.286 | 0.943 |
| 20 Nov | Adjacency | 2 | 3 | 0.203 | 0.771 |
| 22 Nov | Plain | 2 | 3 | 0.315 | 0.829 |
| 22 Nov | Adjacency | 2 | 3 | 0.239 | 0.714 |
| 1 Dec | Plain | 2 | 3 | 0.304 | 0.943 |
| 1 Dec | Adjacency | 2 | 3 | 0.185 | 0.886 |

Note. The target is $\geq 2$ reported cases in the health zone during December 2019. Top-k values count realised-positive zones captured among the k highest priorities. The cumulative-burden comparator captured 2/4 at Top 3 and 3/4 at Top 4 at every analysed date. Brier scores are descriptive because the evaluation contains six zones and one target month.

**Figure 5.**
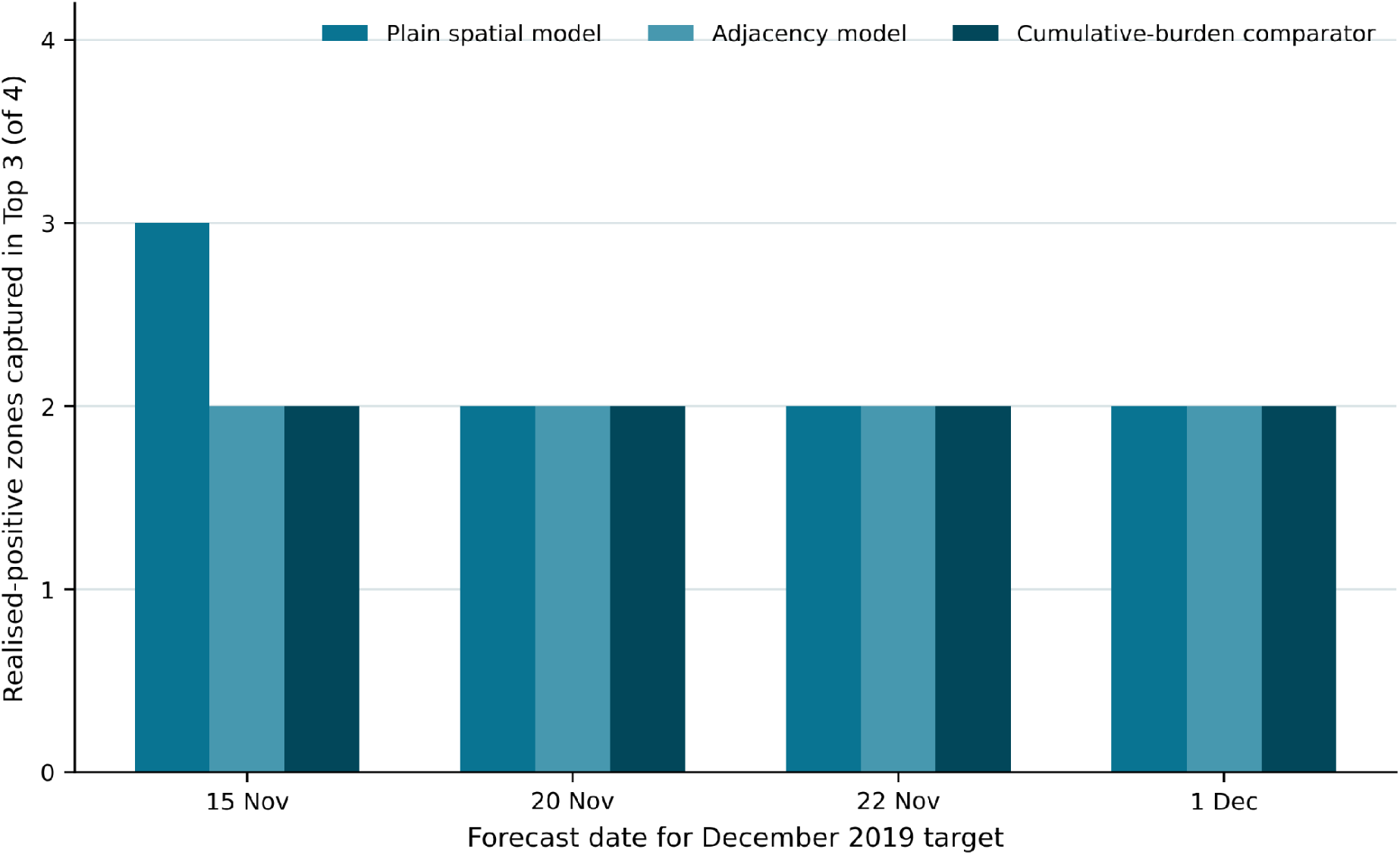
Top-3 geographic prioritisation across the surveillance-shock period. Bars show realised-positive health zones captured among the three highest December priorities. The cumulative-burden comparator captured two of four positive zones at every origin.

The probability and ranking metrics did not move together. The adjacency specification produced a lower mean Brier score than the plain model at all four dates (0.199 versus 0.220; 0.203 versus 0.286; 0.239 versus 0.315; and 0.185 versus 0.304, respectively), yet it never improved Top-3 or Top-4 capture over the cumulative-burden comparator. At several origins it also changed the identity of the missed positive zone by increasing Mambasa risk while assigning an even higher risk to Mandima. A specification could therefore improve a probability score without improving the allocation decision for which the ranking would be used.

These results do not yet justify a Class D designation. Under the predefined framework, Class D requires evidence that a ranking repeatedly outperforms a relevant comparator on the decision-matched metric. Here, only one pre-shock origin of one specification improved the Top-3 decision, and the evaluation contains a single target month. The strongest permissible interpretation is therefore that geographic risk ordering showed continuity through the surveillance disturbance, but validated rank-only readiness was not established. This negative result is itself consistent with the framework: stability of a ranking is not equivalent to demonstrated operational value.

### External validation confirms horizon dependence but rejects a universal M threshold

The preserved Western Area series contained 27 weekly reported-incidence observations, with 24, 23, 22 and 21 evaluable persistence forecasts at one-, two-, three- and four-week horizons, respectively. Point error increased monotonically with forecast horizon. Mean absolute error rose from 39.1 reported cases at one week to 56.2 at two weeks, 69.4 at three weeks and 82.3 at four weeks; mean sAPE increased in parallel from 0.261 to 0.413, 0.532 and 0.635.

The published probabilistic evaluation showed the same horizon gradient. For the best calibrated semi-mechanistic specification, the calibration p-value declined from 0.26 at one week to 0.14 at two weeks and 0.03 at three weeks. Funk et al. reported increasing evidence of miscalibration at three weeks or more and poor calibration for all model variants at four weeks or longer. Mean absolute error of the semi-mechanistic median forecast increased from 42 to 65 and 93 cases across the first three horizons; the corresponding autoregressive values were 43, 60 and 73, and the unfocused values were 47, 61 and 71 [8].

**Table 6.** External forecast validation in Western Area, Sierra Leone.

| Horizon | n | Persistence MAE | Persistence sAPE | Semi-mech calibration | Semi-mech AE | AR AE | Unfocused AE |
| --- | --- | --- | --- | --- | --- | --- | --- |
| 1 week | 24 | 39.1 | 0.261 | 0.26 | 42 | 43 | 47 |
| 2 weeks | 23 | 56.2 | 0.413 | 0.14 | 65 | 60 | 61 |
| 3 weeks | 22 | 69.4 | 0.532 | 0.03 | 93 | 73 | 71 |
| 4 weeks | 21 | 82.3 | 0.635 | Poor for all variants | — | — | — |

Note. Persistence results were reconstructed from the preserved 27-week Western Area incidence series. Published semi-mechanistic and null-model metrics are from Funk et al. [8]. Published absolute-error values are reported for one- to three-week horizons; the paper states that calibration was poor for all model variants at four weeks or longer. AR = autoregressive.

**Figure 6.**
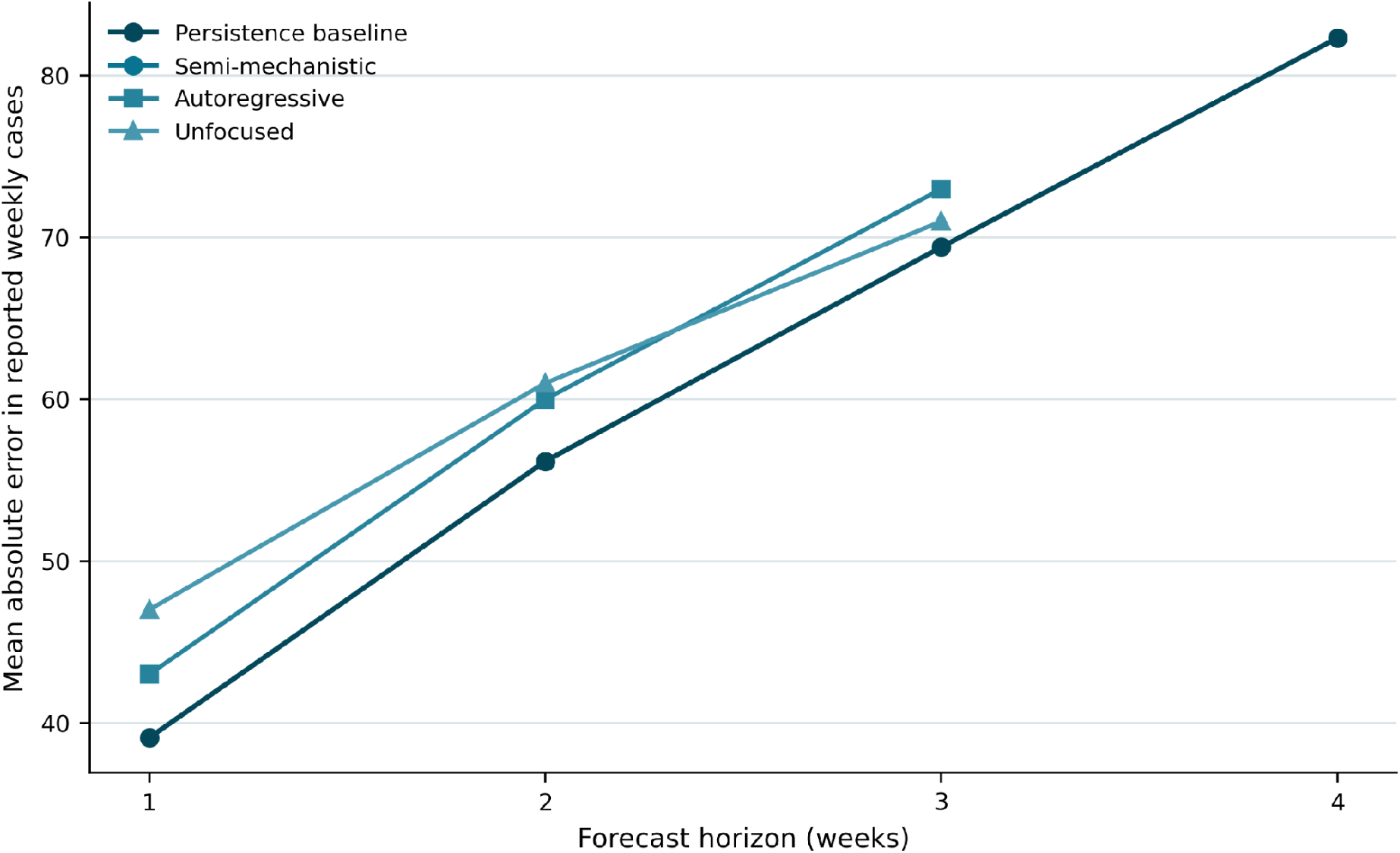
Forecast error increases with horizon in Western Area, Sierra Leone. Published model values are available for one- to three-week horizons; the reconstructed persistence comparator is shown through four weeks.

The persistence comparator is intentionally not a probabilistic substitute for the semi-mechanistic model. Its relevance is that central forecast error was in the same broad range as the published model medians: at one week, persistence MAE was 39.1 compared with 42 for the semi-mechanistic model; at two weeks, 56.2 compared with 65; and at three weeks, 69.4 compared with 93. This does not imply that persistence was the better forecast overall because it provides no calibrated predictive distribution. It does show that additional model complexity did not guarantee lower point error, reinforcing the requirement for explicit baseline comparison.

The history-to-horizon ratio did not behave as a monotonic readiness score. Mean sAPE was 1.025 for the single observation with M<1, 0.670 for 1<=M<2, 0.369 for 2<=M<4, 0.533 for 4<=M<8, and 0.345 for M>=8. These bins confound epidemic phase, horizon and information accumulation and were not used for inferential testing. Their non-monotonic pattern nevertheless provides independent evidence against treating M>=1, or any simple elapsed-history threshold, as sufficient for readiness.

The external validation therefore supports P2 while rejecting a universal history-to-horizon threshold. Forecast horizon is clearly consequential, and history relative to horizon remains a useful diagnostic quantity, but the ratio must be interpreted jointly with observation-regime stability, epidemic-process change and empirical out-of-sample performance. This is also consistent with the original Western Area analysis: model calibration at one or two weeks was comparatively strong from early in the epidemic, whereas longer-horizon calibration remained poor despite the accumulation of additional historical data.

### Precision in reported mortality does not identify latent fatality

The targeted mortality experiment produced structural non-identifiability by construction. Five substantively different latent mechanisms yielded the same expected aggregate observation of 1,000 reported cases and 400 reported deaths (rCFR = 0.40). The implied true fatality ranged from 0.10 under low case ascertainment and complete death ascertainment to 0.80 under either incomplete death ascertainment or incomplete outcome maturity.

**Table 7.** Distinct latent mechanisms with the same expected rCFR.

| Mechanism | N | p | qC | qD | u | Expected C / D / $rCFR$ |
| --- | --- | --- | --- | --- | --- | --- |
| M1 | 4000 | 0.10 | 0.25 | 1.00 | 1.00 | 1000 / 400 / 0.40 |
| M2 | 2000 | 0.20 | 0.50 | 1.00 | 1.00 | 1000 / 400 / 0.40 |
| M3 | 1000 | 0.40 | 1.00 | 1.00 | 1.00 | 1000 / 400 / 0.40 |
| M4 | 1000 | 0.80 | 1.00 | 0.50 | 1.00 | 1000 / 400 / 0.40 |
| M5 | 1000 | 0.80 | 1.00 | 1.00 | 0.50 | 1000 / 400 / 0.40 |

*Note.* $p$ = latent fatality; $qC$ = case ascertainment; $qD$ = death ascertainment; $u$ = fraction of eventual fatal outcomes observable by the analytical origin. These are deliberately simplified identifying examples rather than epidemiological reconstructions of a named outbreak.

The stochastic replicates confirmed that the aggregate observation remains weakly discriminating even when finite-sample variation is included. Across the five mechanisms, mean simulated rCFR remained essentially 0.40, with 95% empirical ranges of approximately 0.36–0.45 for M1, 0.36–0.44 for M2, 0.37–0.43 for M3, 0.37–0.43 for M4 and 0.37–0.43 for M5. Despite true fatality differing eightfold between the lowest and highest mechanisms, their observed-ratio distributions overlapped strongly.

The precision experiment separated sampling uncertainty from identification. With 100 reported cases and an rCFR of 0.40, the naive Wilson 95% interval was approximately 0.31–0.50. At 1,000 reported cases it narrowed to 0.37–0.43; at 10,000 cases to 0.39–0.41; and at 25,000 cases to approximately 0.394–0.406. In contrast, under the predefined ρ = qDu/qC ∈ [0.5, 2] range, the latent fatality interval remained fixed at 0.20–0.80 at every denominator.

**Figure 7.**
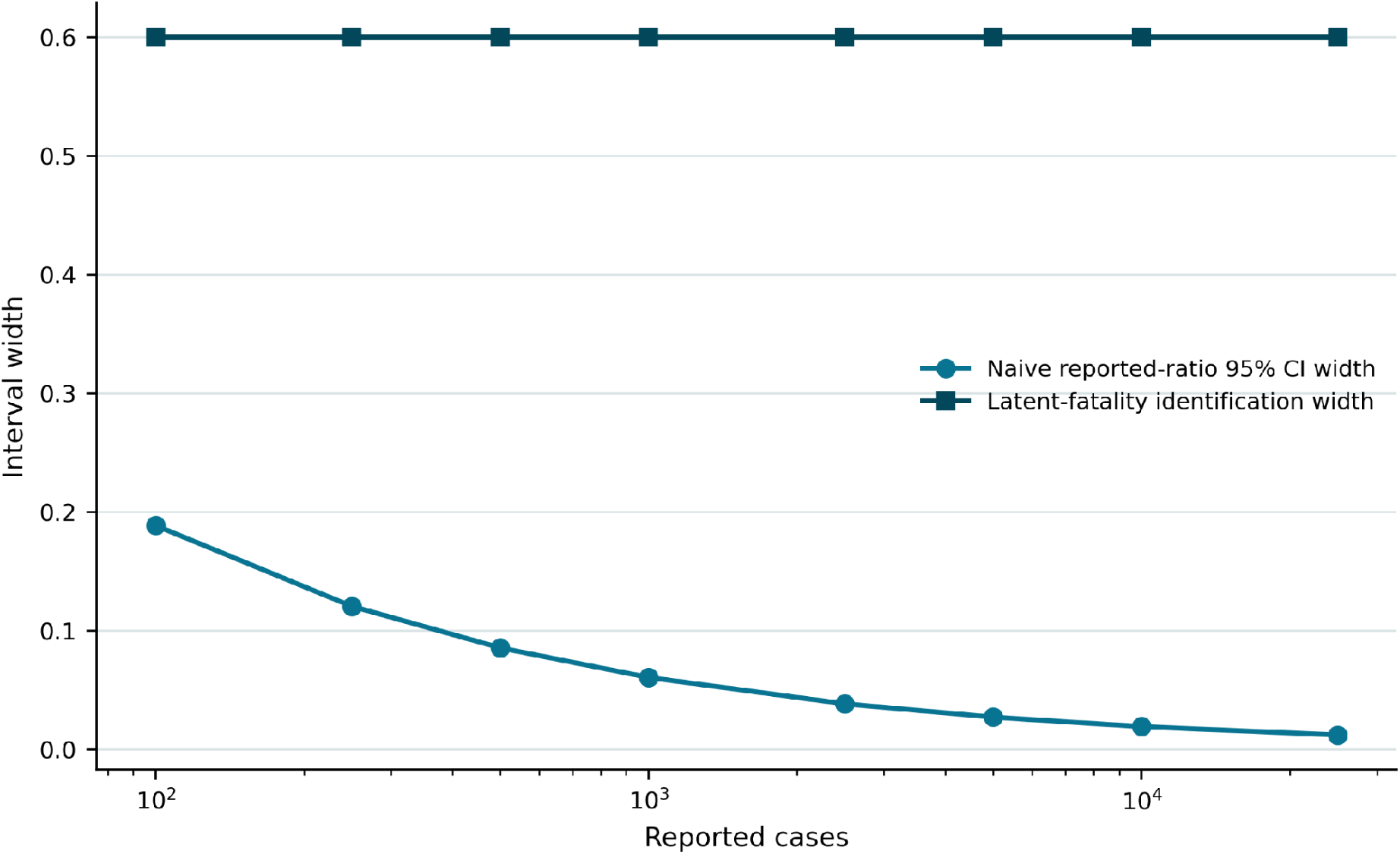
Sampling precision does not resolve latent-fatality identification. Increasing the reported denominator narrows the naive 95% interval around rCFR while the structural identification width remains unchanged under fixed relative-ascertainment bounds.

Allowing incomplete outcome maturity widened the equivalence set further. In the explicit M5 mechanism, latent fatality was 0.80 with complete case and death ascertainment but only half of eventual fatal outcomes observable by the origin; this generated the same expected rCFR as the 0.40 complete-ascertainment benchmark. Thus, a narrow interval around rCFR cannot distinguish low fatality with preferential death ascertainment, high fatality with incomplete death ascertainment, or high fatality with unresolved outcomes.

These findings map directly onto the readiness classes. The reported crude case-fatality ratio is a Class E quantity when used descriptively: it is observable and can be estimated precisely, but precision does not license biological interpretation. Latent fatality p is Class F when qC, qD and u are unrestricted because substantively different latent states remain observationally equivalent. If external evidence supplies defensible bounds on ρ, the estimand can move to Class C and be reported as a scenario interval. Movement to Classes B or A would require additional identifying information such as linked patient outcomes with mature follow-up, independent ascertainment estimates, mortality-completeness evidence or validated outcome-delay distributions. This provides a direct demonstration of P3 and P4.

### Cross-module synthesis

Across modules, readiness behaved as an estimand-specific trajectory rather than a dataset-wide state. Surveillance recovery did not coincide automatically with analytical recovery; forecast and trend readiness differed within the same DRC sequence; greater historical depth did not remove horizon-dependent forecast failure; and a larger mortality denominator increased precision without identifying latent fatality.

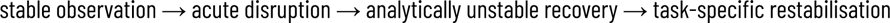

The demonstrations were coherent with the four propositions without implying universal validation: P1 was supported by asynchronous forecast and trend behaviour; P2 by horizon dependence and divergence between Brier and Top-k performance; P3 by overdispersion and mortality equivalence; and P4 by the need for restricted forecasts, rankings, descriptive quantities, scenario ranges or non-identifiability results. No single numerical threshold classified readiness consistently.

## Discussion

### Principal findings

This study operationalises analytical readiness as an estimand- and vintage-specific relationship between evidence and claim. The DRC, Western Area and mortality analyses are structured demonstrations of distinct implications of that framework, not universal proof of one fixed readiness rule. Across tasks, the same information state supported different products, and numerical precision or additional historical depth did not consistently imply stronger inferential support.

### Analytical readiness is estimand-specific, not a property of the dataset

The results support treating analytical readiness as a relationship among the decision, estimand, observation process, method and validation evidence rather than as a permanent attribute of a dataset. This interpretation is consistent with the estimand principle used in other statistical domains, where clarity about the target quantity precedes choice of estimator [10]. It also addresses a common problem in outbreak analytics: methods are often selected because a dataset has reached a certain size, because a model converges, or because a curve appears stable, even though those properties do not establish that the quantity of operational interest has become identifiable or predictively stable.

The DRC results provide a direct within-outbreak example. Forecast readiness, trend-estimation readiness and geographic-prioritisation readiness behaved differently during the same broad surveillance disturbance. The Sierra Leone analysis provides an external analogue: additional historical information did not remove the strong effect of forecast horizon. These findings argue against transferring thresholds mechanically across estimands or outbreaks. A readiness statement should therefore identify the task explicitly—for example, “seven-day reported-burden forecasting is model-dependent at this origin” or “the reported mortality ratio is observable but latent fatality is non-identifiable”—rather than declaring the underlying dataset mature in the abstract.

### Surveillance recovery can be an analytically unstable transition

The DRC surveillance shock suggests that restoration of surveillance activity and restoration of analytical stability need not occur simultaneously. During an acute disruption, the observed series may appear artificially quiet because alerts, contact follow-up, access or reporting are impaired. When surveillance resumes, delayed investigations, renewed case finding, geographic reconciliation and genuine changes in transmission can enter the record together. The resulting report-date series may therefore become more volatile during recovery even as operational indicators improve.

This interpretation should not be overstated causally. The November–December 2019 period also contained real transmission-chain changes, and the available aggregate data cannot decompose the 4–10 December increase into surveillance recovery, delayed recognition and latent epidemic acceleration. The methodological implication is narrower: analysts should not assume that a documented improvement in surveillance process immediately returns all estimands to their pre-disruption readiness class. A recovery phase is itself an observation regime that requires validation.

### Decision-matched validation changes which model is “best”

The spatial analysis illustrates why validation must be tied to intended use. Adjacency-adjusted forecasts had lower Brier error at every evaluated date, yet did not improve the Top-3 or Top-4 allocation decision over the cumulative-burden comparator. Conversely, the plain model improved Top-3 capture at one pre-shock origin but did not sustain that advantage. A ranking can therefore look stable, and a probability model can improve a global scoring rule, without producing a demonstrably better operational priority set.

The same principle applies to temporal forecasting. Proper probabilistic scoring and calibration remain essential where uncertainty distributions are used [5,6], but transparent baselines are also necessary. In Western Area, the simple persistence comparator had central absolute error in the same broad range as the published model medians, even though it did not provide the calibrated predictive distribution required of a full probabilistic forecast. Complexity therefore adds value only relative to the decision and metric being evaluated, not by virtue of complexity itself.

### Precision, model convergence and identification are different evidential claims

The mortality experiment formalises a distinction that is often blurred in real-time outbreak reporting. A narrow interval around a reported deaths-to-cases ratio demonstrates precision about that ratio under its sampling assumptions. It does not establish that the ratio equals a patient-level probability of death. When case ascertainment, death ascertainment and outcome maturity differ, aggregate counts identify a product of latent fatality and relative observation probabilities rather than fatality alone.

Likewise, numerical model convergence is a computational property rather than proof of parameter identification. The O3 Poisson trend fit converged and, under unadjusted model variance, could have appeared to show a fairly precise increase. The large Pearson dispersion visible in the same four observations substantially widened the uncertainty once acknowledged. These results are consistent with the general identifiability literature: apparently well-behaved likelihood or posterior surfaces can conceal practical or structural non-identifiability when multiple parameter combinations remain compatible with the observations [11]. Explicit Class F conclusions should therefore be regarded as informative scientific results rather than analytical failures.

### Relationship to surveillance maturity and real-time forecasting practice

The framework complements rather than replaces conventional surveillance evaluation. Attributes such as timeliness, completeness, representativeness and stability remain properties of the surveillance system [3]. Analytical readiness asks a subsequent question: given those observation properties and the epidemic information accumulated, what inference is supportable now? Separating these layers avoids circular definitions in which a smooth epidemic curve is taken as proof that the surveillance system is mature and then used again as the basis for unqualified inference.

The external Sierra Leone validation also cautions against reducing forecast readiness to a history-to-horizon threshold. The ratio M=H/h remains intuitively useful because long horizons demand more information than short horizons, but the non-monotonic M-bin results show that elapsed history is confounded with epidemic phase and observation regime. The stronger and more transportable formulation is therefore conditional: forecast horizon should be evaluated relative to usable historical information, but readiness must ultimately be demonstrated through time-respecting out-of-sample performance rather than inferred from M alone.

### Operational implications

For real-time analysis teams, the framework suggests a short sequence of questions before a result is operationalised. What decision is being supported? What exact quantity would answer that decision? How does the surveillance system transform the latent event into the available observation? What information has accumulated for that estimand? What validation is possible at the current origin? And what is the strongest interpretation supported by that evidence? These questions do not require a new statistical model. In many settings they instead justify a simpler model, a shorter horizon, a ranked rather than probabilistic product, a scenario range, or an explicit statement that the requested biological quantity is not identifiable.

The inferential-support classes discipline interpretation rather than rank datasets. Class A and B support numerical estimation with different robustness; Class C preserves explicit scenarios; Class D supports a validated ranking without calibrated probabilities; Class E protects directly observed surveillance quantities from biological over-interpretation; and Class F records non-identifiability. Moving from one class to another is a change in the kind of support available, not necessarily an improvement on a single scale.

### Transportability and modular use

The framework is intended to be portable at the level of analytical architecture, not at the level of fixed indicators or thresholds. S must be characterised using the observation system relevant to the setting—for example laboratory and investigation processes in an outbreak response, or completeness, timeliness and reporting-unit participation in a routine DHIS2 architecture. E likewise depends on the estimand: the information required for a one-week burden forecast differs from that required for a clinical fatality estimate or geographic-prioritisation decision.

The DRC and Western Area analyses therefore demonstrate how the framework can distinguish readiness across tasks and time in two Ebola surveillance systems; they do not establish that the same process indicators, horizons or empirical cut-offs should be transferred unchanged to cholera, measles, influenza or other surveillance architectures. Transportability should be assessed by preserving the workflow—decision, estimand, observation process, S, E, method, validation and permissible inference—while adapting the task-specific indicators and validation standards.

The modular design is a practical strength in this respect. A response team can apply only the module relevant to its decision: forecast readiness without a spatial model, prioritisation without latent-fatality inference, or descriptive Class E reporting where stronger identification is impossible. Generalisability should therefore be evaluated module by module rather than by requiring every outbreak to reproduce the full set of demonstrations used here.

### Limitations

This study has several limitations. First, the DRC surveillance shock is a quasi-experimental stress test rather than a randomized or instrumental-variable design. Security disruption affected surveillance operations but may also have changed transmission, care seeking and response behaviour; causal effects of surveillance quality cannot therefore be isolated. Second, the process-side indicators are available at heterogeneous geographic and temporal resolutions and cannot be combined into a validated scalar maturity score. That heterogeneity is part of the framework’s motivation but limits formal effect estimation.

Third, the archived spatial analysis contains six surveyed health zones and one target month. Its negative Class D conclusion is therefore appropriately conservative and should not be read as a general assessment of the underlying spatial model. Fourth, the Western Area external validation uses a simple reconstructed persistence baseline alongside published model metrics rather than re-estimating the full semi-mechanistic model. This is sufficient for testing horizon dependence and baseline relevance but not for claiming a new model comparison. Fifth, the mortality experiment is deliberately simplified. It demonstrates an identification problem under transparent ascertainment and outcome-maturity parameters; real case-fatality inference additionally involves correlated ascertainment, delays, case definitions, competing risks and outcome linkage.

Finally, the inferential-support classes have not yet been prospectively evaluated as operational decision rules across many pathogens, surveillance architectures or decision settings. The present study deliberately avoids universal numerical cut-offs, and its empirical evidence is concentrated in Ebola surveillance systems. The framework should therefore be read as operationalised and stress-tested, not as a universal scoring instrument. Future work should test prospectively recorded classifications, additional pathogens and routine-surveillance architectures, and inter-analyst reproducibility of the class assignment rules.

## Conclusions

Real-time outbreak analysis does not become valid at a single moment when a dataset crosses an implicit threshold of maturity. Readiness is estimand- and vintage-specific: the same information state can justify a numerical forecast, a ranking, a descriptive quantity, a scenario range or an explicit conclusion of non-identifiability. The DRC, Western Area and mortality demonstrations show why observation-system quality, epidemic signal, horizon, estimator, validation metric and intended decision must be considered together.

The practical consequence is simple but demanding: define the decision and estimand first; treat surveillance data as outputs of an observation process; characterise S and E; validate the chosen method in a time-respecting and decision-matched way; and report only the strongest inference that evidence supports. More data, more complex models and narrower intervals are useful only when they increase information about the quantity that matters. Analytical readiness is therefore not a property of the dataset, but a defensible relationship between evidence and claim.

## Data Availability

All data produced in the present study are available upon reasonable request to the authors

## List of abbreviations

A–F: inferential-support classes
AE: absolute error
BVD: Bundibugyo virus disease
DRC: Democratic Republic of the Congo
E: epidemic-process informativeness
H: usable historical information
M: history-to-horizon ratio
rCFR: reported crude case-fatality ratio
Rθ,v: estimand-specific analytical readiness for estimand θ at data vintage v
S: surveillance maturity
sAPE: symmetric absolute percentage error
WHO: World Health Organization.

## Declarations

### Ethics approval and consent to participate

This methodological study used publicly available, aggregated and de-identified outbreak surveillance information and public historical archives and did not involve recruitment, intervention, or access to identifiable individual-level records.

### Consent for publication

Not applicable. The manuscript contains no individual person’s data, images or videos.

### Availability of data and materials

The datasets supporting the conclusions of this article are included within the article and its additional files. Additional file 2 contains compact analysis-ready inputs, source lineage and reproducible Python and R code; public upstream source locations are recorded in the source manifest. The redistributed inputs reproduce the manuscript calculations but do not replace the originating surveillance repositories.

### Authors’ contributions

JGLV conceived the study, developed the framework and methodology, curated the analytical data, performed the formal analyses, developed the software and reproducibility materials, prepared the visualisations, and drafted the manuscript. CNM and WJ reviewed the methodology and interpretation and contributed to critical revision of the manuscript. All authors read and approved the final manuscript.

## Acknowledgements

Not applicable.

## Additional files

Additional file 1. File format: DOCX. Title: Supporting Information S1–S8. Description: Extended framework definitions and operational indicators; estimand–evidence–validation matrices; provenance and surveillance-shock reconstruction; detailed DRC, spatial, Western Area and mortality methods; reproducibility and application guidance.

Additional file 2. File format: ZIP. Title: Analytical readiness reproducibility archive. Description: Compact analysis-ready inputs, source manifest, Python and R scripts, expected outputs, figure-generation scripts and checksums for the analyses reported in the manuscript.

